# Temporal changes in the positivity rate of common enteric viruses among paediatric admissions in coastal Kenya, in the period spanning the COVID-19 pandemic, 2019-2022

**DOI:** 10.1101/2023.07.24.23293059

**Authors:** Arnold W. Lambisia, Nickson Murunga, Martin Mutunga, Robinson Cheruiyot, Grace Maina, Timothy O. Makori, D. James Nokes, Charles N. Agoti

**Author notes:** Corresponding author: Contact details: P.O. Box 230, Kilifi-80108, Kenya.

## Abstract

**Background:** The non-pharmaceutical interventions (NPIs) implemented to curb the spread of SARS-CoV-2 early in the COVID-19 pandemic years, disrupted the activity of other respiratory viruses. There is limited data from low-and-middle income countries (LMICs) to determine whether COVID-19 NPIs also impacted the epidemiology of enteric viruses. We investigated the changes in infection patterns of common enteric viruses among hospitalised children who presented with diarrhoea to a referral hospital in coastal Kenya, in the period spanning the COVID-19 pandemic.

**Methods:** A total of 870 stool samples from children under 13 years of age admitted to Kilifi County Hospital between January 2019, and December 2022 were screened for rotavirus group A (RVA), norovirus genogroup II (GII), astrovirus, sapovirus, and adenovirus type F40/41 using real-time reverse-transcription polymerase chain reaction. The proportions positive across the four years were compared using the chi-squared test statistic.

**Results:** One or more of the five virus targets were detected in 282 (32.4%) cases. A reduction in the positivity rate of RVA cases was observed from 2019 (12.1%, 95% confidence interval (CI) 8.7% - 16.2%) to 2020 (1.7%, 95% CI 0.2% – 6.0%; p *< 0.001*). However, in the 2022, RVA positivity rate rebounded to 23.5% (95% CI 18.2% - 29.4%). For norovirus GII, the positivity rate fluctuated over the four years with its highest positivity rate observed in 2020 (16.2%; 95% C.I, 10.0% – 24.1%). No astrovirus cases were detected in 2020 and 2021, but the positivity rate in 2022 was similar to that in 2019 (3.1% (95% CI 1.5% - 5.7%) vs 3.3% (95% CI 1.4% – 6.5%)). A higher case fatality rate was observed in 2021 (9.0%) compared to the 2019 (3.2%), 2020 (6.8%) and 2022 (2.1%) (*p <0.001*).

**Conclusion:** Our study finds that in 2020 the transmission of common enteric viruses, especially RVA and astrovirus, in Kilifi Kenya may have been disrupted due to the COVID-19 NPIs. After 2020, local enteric virus transmission patterns appeared to return to pre-pandemic levels coinciding with the removal of most of the government COVID-19 NPIs.

## Introduction

Although water sanitation and hygiene (WASH) programmes and new vaccine introductions have resulted in significant reductions of paediatric diarrhoea morbidity and mortality globally, virus-associated diarrhoea is still a major cause of hospital admissions in several low and middle-income settings ^1^. In 2019, approximately 300,000 deaths were recorded globally in children below 14 years of age due to rotavirus group A (RVA), norovirus GI and GII, and adenovirus 40/41 infections ^2^.

Following the emergence of COVID-19, a number of reports have indicated perturbations in in seasonality, prevalence and incidence of common enteric viruses associated with diarrhoea disease. For instance, in France ^3^, Poland ^4^, China ^5, 6^ and USA ^7^, the prevalence of RVA during the year 2020 was lower compared to 2018 and 2019. However, in 2021 there was a surge of RVA cases in these countries. Like RVA, a decrease in cases of norovirus GII, sapovirus, adenovirus 40/41 and astrovirus was reported in 2020 in Spain ^8^ and Korea ^9^ compared to previous years. However, this decrease in virus detection has been followed by remarkable outbreaks in 2021 ^5, 10^. Sporadic outbreaks of norovirus have also been reported in China in September 2020 and in the USA, where a total of 992 norovirus outbreaks were reported between August 2021 and July 2022 ^5, 11, 12^. The decline of the detection rates of enteric viruses in the early COVID-19 pandemic phase has been postulated to be a result of the stringent non-pharmaceutical interventions (NPIs) that were implemented to abrogate the pandemic ^13^. Some of the measures included those that may impact enteric pathogen transmission such as frequent hand washing, increased hygiene, social distancing, closure of restaurants and restricted movement either locally or internationally ^13^.

In coastal Kenya, the prevalence of enteric viruses over the past decade has been monitored and no significant change in the prevalence has been detected in all the viruses except for sapovirus (7.6% vs 4.0%, *p value* <0.05) pre-post rotavirus vaccine introduction in July 2014 ^14^. RVA positivity in hospital admissions decreased significantly only among ELISA detected cases but not RT-PCR detected cases ^15^. Continuous monitoring of these enteric viruses that commonly cause diarrhoea in childhood is key in providing insights on their epidemiology for disease management and informing public health policy. In this study, we aimed to describe the epidemiological patterns of common enteric viruses associated with diarrhoea between January 2019 and December 2022, during a period spanning the COVID-19 pandemic.

## Methods

### Study site and population

This study was undertaken as part of our routine surveillance of RVA in Kilifi County Hospital (KCH), Kenya ^14, 16, 17^. To be recruited, a participant had to satisfy the following criteria: (a) admitted with diarrhoea as one of their illness symptom(s), (b) aged < 13-year-old, (c) consent given from a parent or guardian to be in the study ^14, 16, 17^ The surveillance started in 2009 and has continued to date (2023). In this analysis we focused on participants recruited between 1^st^ January 2019 and 31^st^ December 2022.

### Laboratory methods

#### Molecular testing for common enteric viruses

##### Total Nucleic Acid (TNA) Extraction

TNA was extracted from 0.2 grams of stool (or 200ul if liquid) using the QIAamp Fast DNA Stool Mini kit (Qiagen, Manchester, UK) and eluted in 200ul of elution buffer as previously described^14, 16^.

##### Virus (RT)-PCR Screening

The extracted TNA was combined with the TaqMan Fast Virus 1-step master mix and virus specific primers (**supplementary table 1**) for each of the five viruses ^16, 18^. and processed on a real-time Quantistudio 5-flex instrument. The reaction mix comprised 2.5μl of the TaqMan master mix, 1.2μl of the primer-probe mix, 3.8μl of nuclease free water and 5μl of TNA. The thermocycling conditions were as follows; 95°C for 20 sec and 35 cycles of 94°C for 15 sec and 60°C for 30 sec. A cycle threshold cut-off of < 35.0 was applied to determine virus positive samples for all targets screened.

### RVA genotyping

TNA from RVA positives were amplified using segment specific primers, sequenced on the Illumina Miseq platform as previously described ^19^. Genomes were assembled from the short read data using a de novo assembly approach as previously described ^19^. RVA genotypes were assigned using either BLAST or an online RVA genotyping tool ^20^.

### Statistical Analysis

All statistical analysis was undertaken using R version 4.1.1 (2021-08-10). The level of government intervention was summarised using the Oxford Stringency index (SI), a composite measure based on nine response indicators including school closures, workplace closures, and travel bans, rescaled to a value from 0 to 100 (100 = strictest) ^21^. Local stringency measures have been highlighted elsewhere ^22^ and summarized in supplementary table 2.

The virus positivity rate during each year was calculated as the proportion of samples that tested positive for the given virus given the total number of samples tested in the defined year. The data from 2019 has been previously published elsewhere and formed a reference base of the situation before COVID-19 ^16^. Comparisons across different years and groups were done using the chi-squared test statistic. Kruskal Wallis and Wilcoxon rank-sum tests were used to compare the distribution of continuous variables. Disease severity was estimated using the Vesikari Clinical Severity Scoring System Manual as previously described ^16, 23^.

## Results

### Basic demographic characteristics

Between January 2019 and December 2022, 1,613 patients aged under 13 years presented with diarrhoea as one of their illness symptoms at KCH. Of these, 870 (54.0%) consented enrolment into the study, gave a stool sample, and were included in this analysis. The reasons for missed sample collection in the study were: consent refusal (n=344), other (n=133), death (n=68), discharged before sample collection(n=35) and transferred before sample collection (n=3).

All the 870 stool samples were screened for the five enteric viruses namely, RVA, norovirus GII, astrovirus, sapovirus and adenovirus F40/41. The majority of the recruited patients were in their first year of life (n=371, 42.6%) and all suffered moderate-to-severe diarrhoeal disease (**Table 1**). The characteristics of the observed cases across the four years in terms of gender and age were similar (*p value > 0.05*). However, fatal outcome appeared more likely to occur in 2021 (9.0%) compared to 2019 (3.2%), 2020 (6.8%) and 2022 (2.1%) (*p value* < 0.001, **Table 1**). Less severe disease was also reported in 2020 compared to the other three years, **Table 1**.

**Table 1:** Demographic characteristics of children under 13 years admitted to Kilifi County Hospital, Kenya between January 2019 and December 2022.

|  | 2019<br>(n=314) * | 2020<br>(n=117) | 2021<br>(n=201) | 2022<br>(n=238) | Total<br>(n=870) | P value |
| --- | --- | --- | --- | --- | --- | --- |
| **Sex** |  |  |  |  |  | 0.586 |
| Female | 137 (43.6%) | 45 (38.5%) | 81 (40.3%) | 107 (45.0%) | 370 (42.5%) |  |
| Male | 177 (56.4%) | 72 (61.5%) | 120 (59.7%) | 131 (55.0%) | 500 (57.5%) |  |
| **Median age in months**<br>**(Interquartile range)** | 14.0 (8.0 – 25.0) | 13.7 (8.1 – 27.0) | 13.0 (7.5 – 23.4) | 13.2 (8.5 – 21.3) | 13.7 (8.1 – 23.0) |  |
| **Age strata (months)** |  |  |  |  |  | 0.252 |
| <12 | 128 (40.8%) | 51 (43.6%) | 89 (44.3%) | 103 (43.3%) | 371 (42.6%) |  |
| 12 -23 | 102 (32.5%) | 36 (30.8%) | 64 (31.8%) | 86 (36.1%) | 288 (33.1%) |  |
| 24 -59 | 56 (17.8%) | 13 (11.1%) | 33 (16.4%) | 35 (14.7%) | 137 (15.7%) |  |
| >60 | 28 (8.9%) | 17 (14.5%) | 15 (7.5%) | 14 (5.9%) | 74 (8.5%) |  |
| **Outcome** |  |  |  |  |  | <0.001 |
| Alive | 276 (87.9%) | 109 (93.2%) | 183 (91.0%) | 231 (97.1%) | 799 (91.8%) |  |
| Dead | 10 (3.2%) | 8 (6.8%) | 18 (9.0%) | 5 (2.1%) | 41 (4.7%) |  |
| Data missing | 28 (8.9%) | 0 (0.0%) | 0 (0.0%) | 2 (0.8%) | 30 (3.4%) |  |
| **Disease severity** |  |  |  |  |  | 0.01 |
| Moderate | 90 (28.7%) | 47 (40.2%) | 45 (22.4%) | 65 (27.3%) | 247 (28.4%) |  |
| Severe | 224 (71.3%) | 70 (59.8%) | 156 (77.6%) | 173 (72.7%) | 623 (71.6%) |  |
| *Pre-pandemic data has been reported in an earlier manuscript <sup>16</sup> |  |  |  |  |  |  |

### Trends in diarrhoeal cases in the context of the COVID-19 pandemic

After the initial detection of the first COVID-19 case in Kenya on 12^th^ March 2020 ^24^, the government implemented a range of NPIs to curb the pandemic (summarised in Figure 1a using the Oxford Stringency index and local restrictions listed in supplementary table 2). The trend of monthly recorded diarrhoea cases recruited into our surveillance between January 2019 and December 2022 is shown in **Figure 1b**. The highest monthly peak in cases was recorded in 2019 before the COVID-19 pandemic. The lowest number of diarrhoea cases was in the 2020, but a gradual rebound was observed in 2021 and 2022.

**Figure 1:**
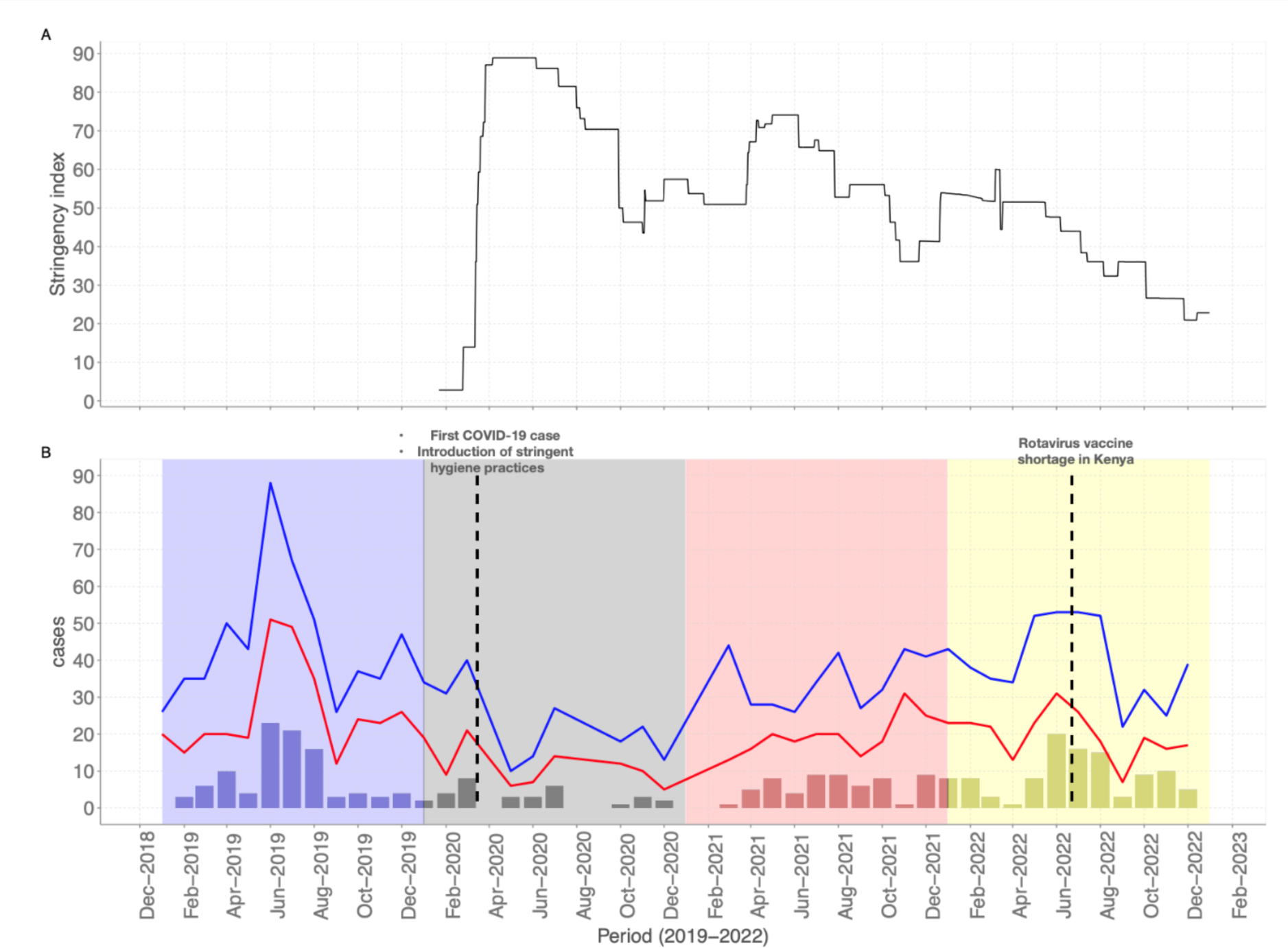
**A)** COVID-19 stringency index quantifying the government NPI measures aimed to curb the spread of SARS-CoV-2 (source: https://ourworldindata.org/covid-stringency-index). B) Temporal trends of monthly diarrhoea and virus cases between January 2019 and December 2022. The blue and red line graphs show the total eligible and recruited diarrhoea cases respectively, while the bar graphs show the total virus positive cases per month.

### Single virus infections and coinfections

At least one of the five virus targets were detected in 282 (32.4%) cases. The proportion of samples positive for the five viruses we tested in the stool samples for the different years is summarised in table 1. The positivity rate of RVA deeped in 2020, at 1.7% (95% C.I, 0.2% – 6.0%) compared to 2019 (12.1% (95% C.I, 8.7%-14.9%) and gradually rose in 2021 (16.9% (95% C.I, 12.0% - 22.8%)) and 2022 (23.5% (95% C.I, 18.2% - 29.4%)) and the differences were statistically significant (p value < 0.001) (**Table 2**).

**Table 2:** Comparison of the detection rates of five common enteric virus between January 2019 and December 2022.

|  | Total (n=870) | 2019 (n=314) |  | 2020 (n=117) |  | 2021 (n=201) |  | 2022 (n=238) |  | P value |
| --- | --- | --- | --- | --- | --- | --- | --- | --- | --- | --- |
|  |  | Cases | Proportion (95% CI) | Cases | Proportion (95% CI) | Cases | Proportion (95% CI) | Cases | Proportion (95% CI) |  |
| **Virus** |  |  |  |  |  |  |  |  |  |  |
| Rotavirus | 130 (14.9%) | 38 | 12.1 (8.7 - 16.2) | 2 | 1.7 (0.2 – 6.0) | 34 | 16.9 (12.0 – 22.8) | 56 | 23.5 (18.2 – 29.4) | <0.001 |
| Norovirus GII | 89 (10.2%) | 34 | 10.8 (7.6 – 14.8) | 19 | 16.2 (10.0 – 24.1) | 13 | 6.4 (3.4 - 10.8) | 23 | 9.6 (6.2 – 14.1) | 0.04 |
| Astrovirus | 18 (2.1%) | 10 | 3.1 (1.5 – 5.7) | 0 | - | 0 | - | 8 | 3.3 (1.4 – 6.5) | 0.01 |
| Sapovirus | 33 (3.8%) | 11 | 3.5 (1.7 – 6.1) | 3 | 2.5 (0.5 – 7.3) | 7 | 3.4 (1.4 – 7.0) | 12 | 5.0 (2.6 – 8.6) | 0.65 |
| Adenovirus F40/41 | 30 (3.4%) | 9 | 2.8 (1.3– 5.3) | 8 | 6.8 (2.9 - 13.0) | 6 | 2.9 (1.1 – 6.3) | 7 | 2.9 (1.1 – 5.9) | 0.19 |

The norovirus GII positivity rate fluctuated over the four years and the difference was statistically significant (p value 0.04) (**Table 2**). The highest positivity rate for norovirus GII was 16.2% (95% C.I, 10.0% – 24.1%) in 2020 and lowest at 6.4% (95% C.I, 3.4% – 10.8%) in 2021 (**Table 2**). No astrovirus cases were detected in the 2020 and 2021, but the cases in 2022 (3.3%) with a similar positivity rate to what was reported in 2019 (3.1%, **Table 2**). The positivity rates for sapovirus and adenovirus type F40/41 did not change across the three phases (*p value* > 0.05, χ2).

Between 2019 and 2022, 13 samples had a coinfection of two or more viruses of the screened viruses. The most common coinfections were RVA and norovirus GII (n=4), adenovirus 40/41 and astrovirus (n=3), and adenovirus 40/41 and sapovirus (n=3) (**Supplementary table 3**).

### Monthly virus trends

In all the years, peak RVA cases were observed in the month of August, except in 2020 where only two RVA cases were detected, **Figure 2**. The peak months for norovirus GII varied across the different years. Less than five cases were reported over the four years for adenovirus 40/41, sapovirus and astrovirus in each month. In 2020 and 2021, no astrovirus cases were detected but re-emerged in 2022 (**Figure 2**).

**Figure 2:**
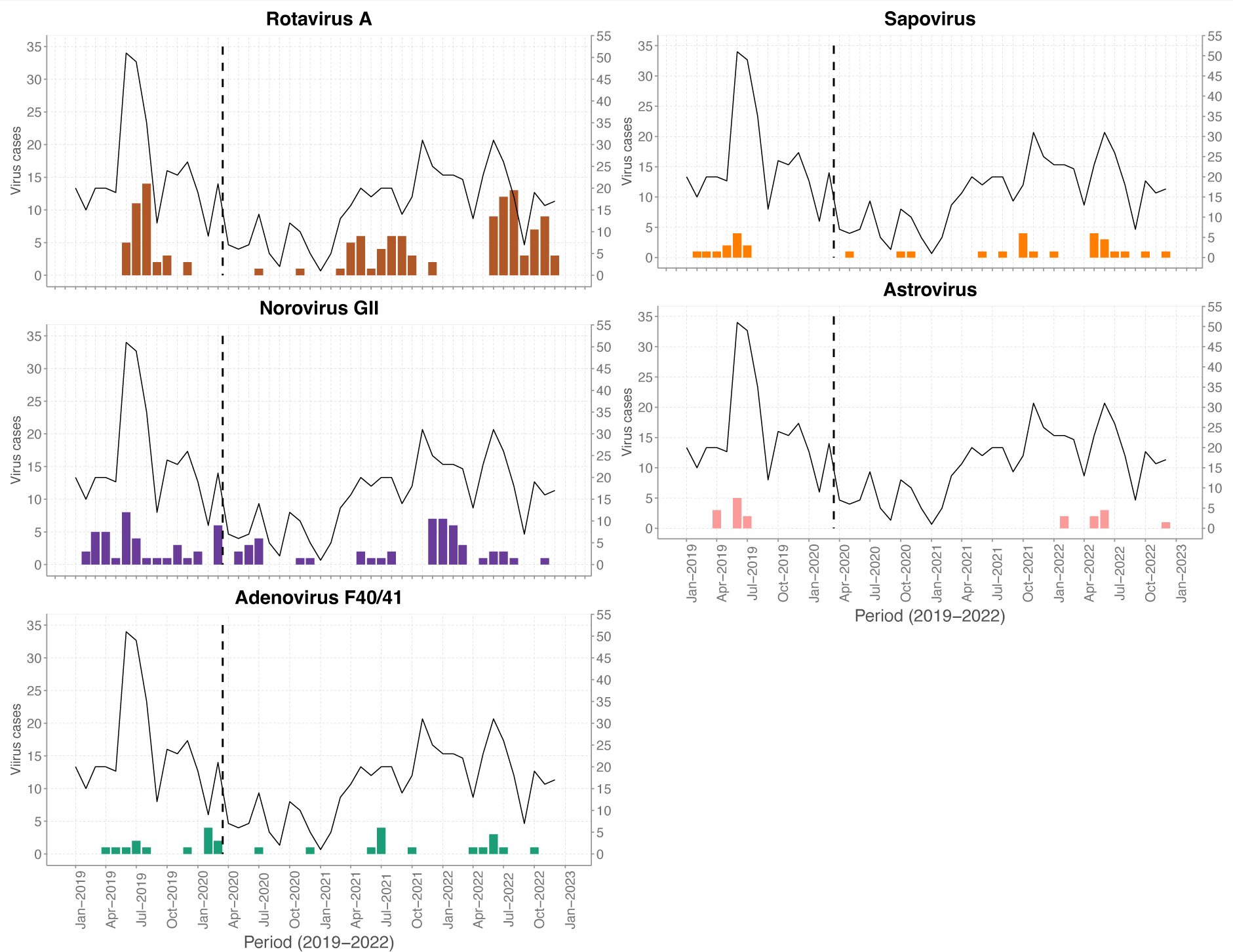
Monthly temporal distribution of common enteric virus cases in children under 13 years admitted to KCH with diarrhoea between January 2019 and December 2022. The black trendline shows the monthly diarrhoeal cases over time and the vertical dotted line represents when the first COVID-19 case was detected in Kenya.

### RVA genotypes and vaccination status of positive cases

Of the 130 RVA positive detected during the study, 87 (66.9%) and 70 (53.8%) successively sequenced in the VP7 and VP4 segment, respectively. The years 2019, 2020 and 2021 were predominated by the G3P[8] genotype (n=48, 80.0%). In 2022, we observed replacement of the G3P[8] genotype with multiple genotypes: G2P[4] (n=2), G9P[8] (n=10), and G9P[4] (n=4). To note, in the 2022 there was some incomplete genotyping due to failed sequencing in the VP4 and VP7 segments G2P[x] (n=5) and GxP[8] (n=8), Among the 130 RVA positive cases, 88 (67.7%) had received at least one dose of the Rotarix® vaccine two doses, 18 (12.9%) had not received a vaccine and 24 (16.9%) had no vaccination records. One participant was RVA positive five days after receiving a Rotarix dose and their sample genotyped as G3P[8].

### Disease outcome

In the study, a total of 41 (4.7%) cases succumbed among the 870 that we analysed. Only nine of these cases were positive for at least of the viruses we tested i.e., norovirus GII (n=4), adenovirus type F40/41 (n=2), RVA (n=1), sapovirus (n=1) and a coinfection (RVA & sapovirus, n=1) (**Supplementary Figure 2).** The other 32 cases had none of the five viruses detected.

## Discussion

In this coastal Kenya hospital-based study, we observed a decrease in paediatric diarrhoea admissions in during the first year of the COVID-19 pandemic (2020) compared to the pre-pandemic year (2019). This decline may be attributed in part to be a result of the implementation of NPIs such as restriction of movement and increased hygiene practices that may have led to reduce transmission of the enteric pathogens as observed elsewhere ^8^ and the overall reduced access to the hospital occasioned by health worker strike between December 2020. Peak diarrhoea cases are usually observed between June and July and in 2020 and 2021, this period coincided with high stringency measures in the country. After March 2021, targeted vaccination campaigns started replacing NPIs and from August 2021, the government dropped measures including closure of schools, curfews, lockdowns and public gatherings and restrictions in public transport.

We report a significant decrease in RVA positivity rate in 2020 compared to 2019 followed by a rebound in 2021 and 2022, a finding consistent with other studies elsewhere ^4, 6–8^. Notably, the positivity rate of RVA in 2022 was higher compared to all the previous years. Such a resurgence of RVA activity was also observed in Hong Kong after the first year of the pandemic^5^.

The return and increase of RVA activity in the 2021 and 2022 can be attributed in part to the relaxation of the COVID-19 NPIs or circulation of strains heterologous to the Rotarix vaccine (e.g. G9P[8], G9P[4] and G2P[4]). Some of these strains have been noted to have limited cross-reactivity with the Rotarix G1P[8] strain. Further, vaccination delays and Rotarix® vaccine stockouts that occurred in between June 2022 and January 2023 when a vaccine switch to Rotavac was made in Kenya ^25^. The introduction of G9 genotypes in Malaysia was associated with an increase RVA prevalence ^26^ similar to what we observed in 2022 whereby the G3P[8] were rapidly replaced by the G9P[8], G9P[4] and G2P[4] genotypes and subsequently there was an increase in RVA cases.

In Kilifi, the prevalence of norovirus GII has been on the rise post-rotavirus vaccine introduction compared to the pre-vaccine period ^14, 16^. Interestingly, the positivity rate of norovirus GII was highest in 2020 phase compared to other years. This implies that norovirus activity was less impacted by the NPI measures which is known to be highly infectious. An increase in norovirus activity and multiple sporadic outbreaks have reported in the USA and China during the COVID-19 phase ^5, 11, 12^.

Notably, astrovirus was not detected in the 2020 and 2021 but re-emerged in the 2022 with a positivity rate similar to what was observed in 2019. Astrovirus and sapovirus detection on the Kenyan coast has been always characterised by very low prevalence (<5%) ^14, 16^.

In China, enteric virus coinfections especially with RVA and norovirus was associated with severe disease ^27^. Similarly in coastal Kenya, a coinfection with either RVA or/and norovirus GII coincided with severe disease. However, its key to note that 71.6% of the participants in the study presented with severe disease. In 2020, there was a significant difference in mortality rate (9.0%) compared to the 2019 (3.2%), 2021 (6.8%) and 2022 (2.1). We hypothesize that the high mortality rate in the 2020 and partly 2021 may have been caused by the challenges with access to the hospitals and health care services as efforts were geared towards management of COVID-19 cases.

This study had several limitations. We did not analyse healthy controls from the same population to adjust the aetiological fraction for asymptomatic carriage in the population. With the small number of detections for some of the virus targets, it hard to confidently infer seasonality without a larger study. There was substantial data missingness e.g., on deaths and almost half of the eligible participants refused consent to be in the study. For a conclusive inference of changes of the incidence of these viruses before during and after the pandemic, a population-based study with a clear population-based denominator is necessary.

In conclusion, our study observed a decrease in total diarrhoea admissions and enteric virus activity during in 2020 and 2021. However, in 2022 an increase in RVA and astrovirus activity is restored to pre-pandemic numbers. Therefore, continuous enteric virus surveillance is key in understanding the temporal changes in positivity rate of these viruses to inform public health policy.

## Data Availability

All the data used can be accessed at the KWTRP Research repository via https://doi.org/10.7910/DVN/EG6MEH.

## Declarations

### Ethics approval and consent to participate

The research protocol for the study was approved at Kenya Medical Research Institute (KEMRI), by the Scientific and Ethics Review Unit (SSC#2861) in Nairobi, Kenya.

## Acknowledgement

We are grateful to the study participants who provided samples and members of the pathogen epidemiology and omics group at KEMRI-Wellcome Trust Programme who did sample collection and laboratory processing. This manuscript was written with the permission of Director KEMRI CGMRC

## Consent for publication

Yes.

## Authors’ contributions

CAN and DJN sourced the study funding. CNA, DJN and AWL designed the study laboratory assay. AWL, MM, TOM, CR, and GM did the laboratory experiments. NM and AWL managed the study data and did the data analysis. AWL and CAN wrote the first manuscript draft. All authors read, revised, and approved the final manuscript.

