## Supplementary table 1 for "Temporal changes in the positivity rate of common enteric viruses among paediatric admissions in coastal Kenya, in the period spanning the COVID-19 pandemic, 2019-2022"

### Supplementary table 1: Primer and probe sequences

| Virus | Strand | Sequence |
|---|---|---|
| **Sapovirus** | Primer-F | CAGGCTCTCGCCACCTAC |
| | Primer-R | CCCTCCATYTCAAACACTAWTTT |
| | Probe | TGGTTCATAGGTGGTRC |
| **Astrovirus** | Primer-F | TCAACGTGTCCGTAAMATTGTCA |
| | Primer-R | GCWGGTTTTGGTCCTGTGA |
| | Probe | CAACTCAGGAAACARG |
| **Rotavirus A** | Primer-F | ACCATCTWCACRTRACCCTCTATGAG |
| | Primer-R | GGTCACATAACGCCCCTATAGC |
| | Probe | AGTTAAAAGCTAACACTGTCAAA |
| **Norovirus GII** | Primer-F | CARGARBCNATGTTYAGRTGGATGAG |
| | Primer-R | TCGACGCCATCTTCATTCACA |
| | Probe | GAGGGSGATCGCRATCT |
| **Adenovirus** | Primer-F | CACTTAATGCTGACACGGGC |
| | Primer-R | ACTGGATAGAGCTAGCGGGC |
| | Probe | TGCACCTCTTGGACTAGT |
