## Supplementary table 2 for "Temporal changes in the positivity rate of common enteric viruses among paediatric admissions in coastal Kenya, in the period spanning the COVID-19 pandemic, 2019-2022"

**Supplementary table 2: Counter measures implanted by the Kenyan government to counter the spread of COVID-19 in Kilifi**

| Years | Measures |
|---|---|
| **2020** | Travel restriction from other countries with reported COVID cases |
| | Ban of social gatherings including churches, weddings, and funerals |
| | Restriction of movement into and out of Kilifi county |
| | Closure of schools |
| | Compulsory wearing of masks in public |
| | Ban of local air travel |
| | Restriction of restaurants opening hours and offer take-away services only |
| | Night curfew (7 p.m. to 5 a.m. then 11 p.m. to 4 a.m.) |
| | Suspension of political gatherings and meetings |
| **2021** | Night curfew |
| | Compulsory wearing of masks in public |
| | COVID-19 vaccination launched |
| | Cessation of all movement by air, rail, and road in disease infected areas |
| | Suspension of political gatherings and meetings |
| **2022** | Compulsory wearing of masks in public (up to March 2022) |
| | Compulsory hand washing (up to March 2022) |
| | Social distancing (up to March 2022) |
