## Supplementary table 3 for "Temporal changes in the positivity rate of common enteric viruses among paediatric admissions in coastal Kenya, in the period spanning the COVID-19 pandemic, 2019-2022"

**Supplementary table 3:** Enteric virus coinfections observed in Kilifi, Kenya between 2019 and 2022

| | **2019** | **2020** | **2021** | **2022** |
|---|---|---|---|---|
| **Coinfection** | | | | |
| Norovirus GII & Adenovirus_40/41 | - | 1 | 1 | - |
| Norovirus GII & Sapovirus | - | 1 | - | - |
| Rotavirus A & Adenovirus 40/41 | - | - | - | 1 |
| Rotavirus A & Astrovirus | - | - | - | 2 |
| Rotavirus A & Norovirus GII | 1 | - | - | 1 |
| Rotavirus A & Sapovirus | 1 | - | 1 | 2 |
| Sapovirus & Astrovirus | 2 | - | - | - |
